# Association of the transthyretin variant V122I with polyneuropathy among individuals of African descent

**DOI:** 10.1101/2020.11.10.20219675

**Authors:** Margaret M. Parker, Scott M. Damrauer, Catherine Tcheandjieu, David Erbe, Emre Aldinc, Philip N. Hawkins, Julian D. Gillmore, Leland E. Hull, Julie A. Lynch, Jacob Joseph, Simina Ticau, Alexander O. Flynn-Carroll, Aimee M. Deaton, Lucas D. Ward, Themistocles L. Assimes, Philip S. Tsao, Kyong-Mi Chang, Daniel J. Rader, Kevin Fitzgerald, Akshay K. Vaishnaw, Gregory Hinkle, Paul Nioi

## Abstract

**Introduction:** Hereditary transthyretin-mediated (hATTR) amyloidosis is an underdiagnosed, progressively debilitating disease caused by mutations in the transthyretin (TTR) gene. The V122I variant, one of the most common pathogenic TTR mutations, is found in 3-4% of Black individuals, and has been associated with cardiomyopathy.

**Methods:** To better understand the phenotypic consequences of carrying V122I, we conducted a phenome-wide association study scanning 427 ICD diagnosis codes for association with this variant in Black participants of the UK Biobank (n= 6,062). Significant associations were tested for replication in the Penn Medicine Biobank (n= 5,737) and the Million Veteran Program (n= 82,382).

**Results:** Our analyses discovered a significant association between V122I and polyneuropathy diagnosis (odds ratio = 6.4, 95% confidence interval [CI] = 2.6 to 15.6, P = 4.2 × 10−5) in the UK Biobank,which was replicated in the Penn Medicine Biobank (p=6.0×10^−3^)) and Million Veteran Program (P= 1.8×10^−4^)). Polyneuropathy prevalence among V122I carriers was 2.1–9.0% across biobanks. The cumulative incidence of common hATTR amyloidosis manifestations (carpal tunnel syndrome, polyneuropathy, cardiomyopathy, heart failure) was significantly enriched in V122I carriers versus non-carriers (hazard ratio = 2.8, 95% CI = 1.7–4.5, P = 2.6 × 10−5) in the UK Biobank;37.4% of V122I carriers having a diagnosis of any one of these manifestations by age 75.

**Conclusions:** Our findings show that, although the V122I variant is known to be associated with cardiomyopathy, carriers are also at significantly increased risk of developing polyneuropathy. These results also emphasize the underdiagnosis of disease in V122I carriers with a significant proportion of subjects showing phenotypic changes consistent with hATTR. Greater understanding of the manifestations associated with V122I is critical for earlier diagnosis and treatment.

## Introduction

Hereditary transthyretin-mediated (hATTR) amyloidosis, also known as ATTRv (v for variant) amyloidosis, is an underdiagnosed, progressively debilitating, and fatal disease caused by mutations in the transthyretin (*TTR*) gene. These pathogenic variants result in misfolding of TTR proteins, which accumulate as amyloid deposits in multiple tissues throughout the body, including the heart, nerves, and gastrointestinal tract [1-3]. Organ involvement and symptoms of hATTR amyloidosis can vary by patient and mutation, but carriers of pathogenic variants have an increased risk of developing life-impacting polyneuropathy, cardiomyopathy, and other symptoms (e.g., carpal tunnel syndrome [CTS]) [4, 5].

One of the most common pathogenic *TTR* variants is the valine-to-isoleucine substitution at position 122 (V122I; p.V142I; rs76992529), which is predominantly found in individuals of West African descent [6], with 3–4% of this population carrying the mutation [7]. The variant was originally identified in cardiac TTR amyloid deposits [8], and subsequent studies demonstrated it as a common cause of heart failure (HF) among elderly African patients [9, 10]. V122I hATTR amyloidosis is substantially underdiagnosed [5], and patients are often at an advanced disease stage when finally diagnosed. This long journey to diagnosis also results in poor outcomes for these patients, with median time from diagnosis to death being 2.6 years [9]. Thus, a better understanding of early hATTR amyloidosis disease manifestations is critical to expedite diagnosis and initiate treatment [11]. Unlike some other common *TTR* mutations, the V122I variant has historically been associated with a predominantly cardiac phenotype. However, a better understanding of the full range of disease manifestations associated with this variant is critical to improve disease recognition and allow maximal benefit from currently available therapies.

The present study assessed the association of the V122I genotype with ICD10 (International Statistical Classification of Diseases and Related Health Problems, 10th Revision) disease diagnoses in the UK Biobank, a large, population-based, prospective cohort study [12]. The UK Biobank study does not recruit participants based on any disease outcome, thus affording the unique opportunity to analyze the V122I variant in a population not influenced by referral bias, allowing better understanding of early disease manifestations and variant penetrance. Significant associations were assessed for replication in the Penn Medicine Biobank and Million Veteran Program.

## Methods

### Study design

This phenome-wide association study and Cox regression of V122I carriers and non-carriers assessed the association between the V122I variant and ICD diagnoses, including common manifestations of hATTR amyloidosis, in three large biobanks (UK Biobank, Penn Medicine Biobank, and the Million Veteran Program). Written informed consent was obtained from all participants.

### Study population

The UK Biobank recruited ∼500,000 participants in England, Wales, and Scotland between 2006 and 2010 [13]. The Penn Medicine Biobank includes over 60,000 participants over age 18 years enrolled through the University of Pennsylvania Health System since 2008. The Million Veteran Program is a large, multiethnic cohort within the U.S. Department of Veterans Affairs which includes more than 825,000 veterans over age 18 years who have been recruited since 2011 across the United States. Additional details on the biobanks used in this study are provided in the Supplementary Materials.

This study included 6062 unrelated Black participants of the UK Biobank, defined based on a combination of self-reported ancestry and genetic principal components (based on the “British white” subset defined in Bycroft et al. [14]). To define the Black population, the R package aberrant [15] (lambda value = 40) was run on genetic principal components 1 to 6 provided by the UK Biobank to isolate the largest cluster of participants while excluding population outliers. Principal component analysis on the remaining subset using PLINK v.2.0 [16] identified the principal components used as covariates to control for population stratification.

The Penn Medicine Biobank analysis utilizes data on 5737 Black individuals of genetically inferred African ancestry that passed quality control as described in detail elsewhere [5]. Additionally, genotyped and imputed genetic information was available for 468,961 multiethnic participants from the Million Veteran Program, among which 87,163 participants (including related individuals) were of HARE-assigned African ancestry [17]. Of these, 82,382 unrelated Black participants were included in the current study [18].

### V122I genotyping

The V122I genotype was directly typed by the UK Biobank on either the UK BiLEVE or the UK Biobank Axiom arrays [19], which share > 95% common content. Genotypic data were imputed to the UK10K haplotype, 1000 Genomes phase 3, and Haplotype Reference Consortium reference panels using SHAPEIT3 [20] for phasing and IMPUTE4 [14] for imputation. The V122I variant was directly genotyped on both arrays (info score = 1) and passed all quality control metrics (Hardy–Weinberg equilibrium *P* = 7.2 × 10^−5^, genotyping call rate = 100%), as detailed by Bycroft et al. [21]. Patients with and without the V122I genotype are referred to as “carriers” and “non-carriers”, respectively. The presence/absence of other *TTR* genotypes was not examined in these populations.

### Statistical analysis

Descriptive statistics are presented as means (standard deviations) for continuous variables and percentages for categoric variables. Continuous and categoric variables were compared between V122I carriers and non-carriers using a t-test and a Pearson’s chi-squared test, respectively. All analyses were conducted using complete cases (i.e., no missing data).

### Phenome-wide association analysis and replication

To identify diagnoses associated with the V122I variant, a phenome-wide association analysis (PHEWAS) was performed on any primary or secondary inpatient ICD10 diagnosis code observed in at least 15 Black participants from the UK Biobank (*n =* 427 ICD10 codes). PHEWAS was performed in PLINK [16] using logistic regression controlling for age, sex, and genetic ancestry via 10 principal components. A Bonferroni-corrected *P* = 1.2 × 10^−4^ was considered statistically significant. Significant associations with ICD9 and/or ICD10 diagnoses were assessed for replication in the Penn Medicine Biobank and Million Veteran Program using logistic regression controlling for age, sex, and genetic ancestry. Additional methodologic details are provided in the Supplementary Materials.

### Assessment of common hATTR amyloidosis manifestations in V122I carriers

A variable was created to indicate if UK Biobank participants had been diagnosed with at least one of four common hATTR amyloidosis manifestations, independent of whether the participants had a diagnosis of hATTR amyloidosis, using the following ICD10 codes: polyneuropathy (“G62”), CTS (“G560”), cardiomyopathy (“I42”), and HF (“I50” or “I098”). The association between the V122I genotype and a diagnosis of a common hATTR amyloidosis manifestation was tested using logistic regression and Cox proportional hazards regression controlling for age, sex, and genetic ancestry; *P* < 0.05 was considered statistically significant. Kaplan–Meier curves were used to estimate cumulative incidence of common hATTR amyloidosis manifestations by V122I genotype (additional methodologic details in the Supplementary Materials).

### Characteristics of V122I carriers with common hATTR amyloidosis manifestations

V122I carriers with at least one common hATTR amyloidosis manifestation were compared with V122I carriers without manifestations using logistic regression, controlling for baseline age, sex, body mass index, cigarette smoking status, and genetic principal components.

### Population attributable risk of common hATTR amyloidosis manifestations to the V122I genotype

The population risk of common hATTR amyloidosis manifestations attributable to the V122I genotype was calculated by multiplying the fraction of all diagnoses in V122I carriers by the risk difference between carriers and non-carriers. Risk was estimated using the cumulative incidence of the manifestations at age 75 for V122I carriers and non-carriers (additional methodologic details are described in the Supplementary Materials).

## Results

### Baseline characteristics

A total of 387 of 487,327 genotyped UK Biobank participants carried the V122I variant, of which 80.6% were of self-reported African descent (Supplementary Fig. 1). Analyses were performed in the unrelated Black UK Biobank population (*n =* 6062), which included 240 heterozygotes and three homozygotes for the V122I variant (minor allele frequency = 2.0% or 1 in 50). Characteristics of this population are listed in Table 1. On average, participants were followed up for 7.6 years (range: 0.22–10.0 years) and were 59.6 years old at censoring (range: 42.2–79.0 years) (Supplementary Fig. 2).

**Table 1.**
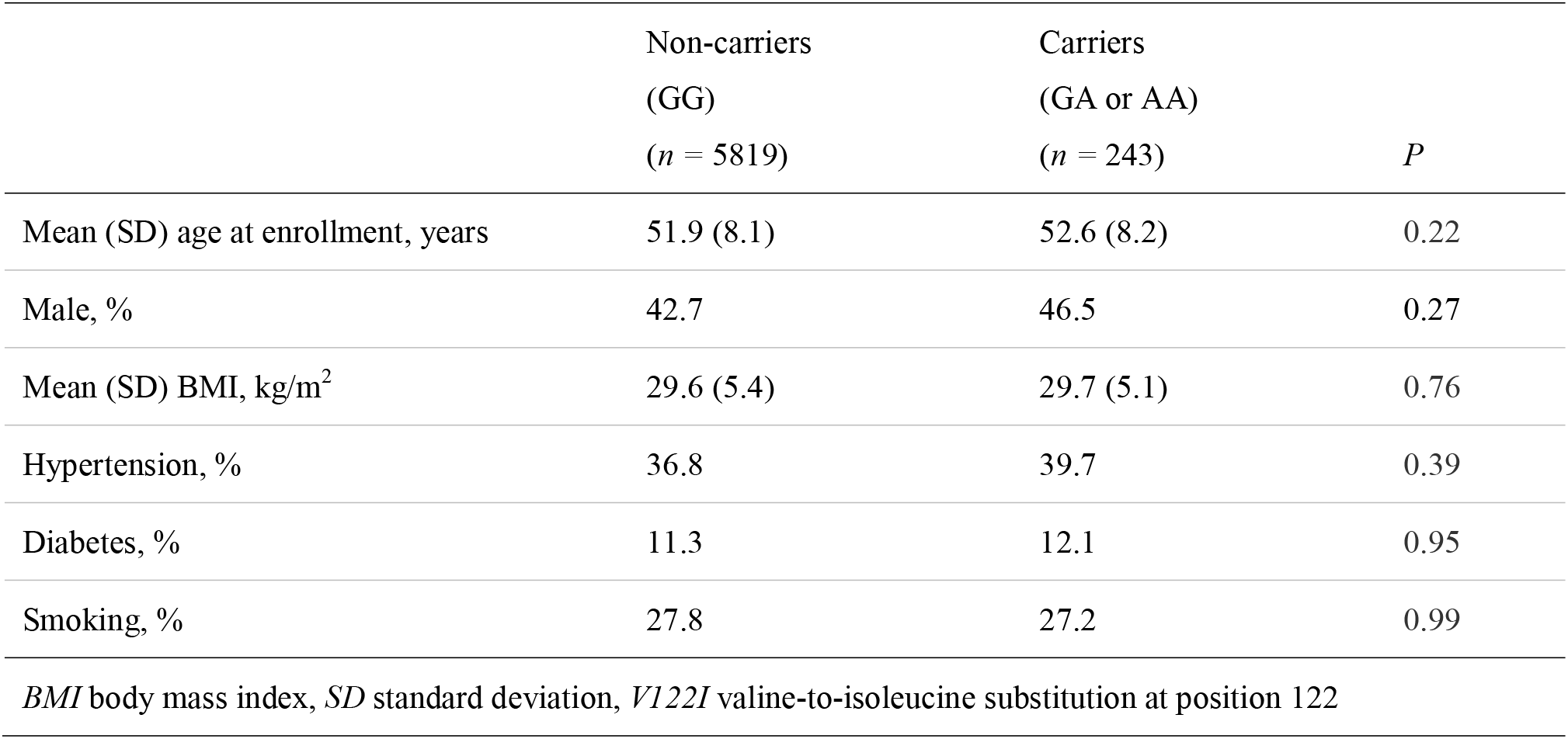
Baseline characteristics of the Black UK Biobank study population (*n =* 6062) by V122I genotype

|  | Non-carriers<br>(GG)<br>(*n* = 5819) | Carriers<br>(GA or AA)<br>(*n* = 243) | *P* |
| --- | --- | --- | --- |
| Mean (SD) age at enrollment, years | 51.9 (8.1) | 52.6 (8.2) | 0.22 |
| Male, % | 42.7 | 46.5 | 0.27 |
| Mean (SD) BMI, kg/m<sup>2</sup> | 29.6 (5.4) | 29.7 (5.1) | 0.76 |
| Hypertension, % | 36.8 | 39.7 | 0.39 |
| Diabetes, % | 11.3 | 12.1 | 0.95 |
| Smoking, % | 27.8 | 27.2 | 0.99 |

*BMI* body mass index, *SD* standard deviation, *V122I* valine-to-isoleucine substitution at position 122

Among the 243 V122I carriers, 0.8% (*n =* 2) were formally diagnosed with amyloidosis (ICD10 diagnosis “E85”) compared with 0.1% (*n =* 5) of non-carriers. Both diagnosed patients were male heterozygous carriers, diagnosed at ages 75.9 and 71.9 years, and with cardiac manifestations of disease diagnosed during study follow-up. One of the two subjects died during study follow-up, with the primary cause of death being organ-limited amyloidosis (ICD10 diagnosis “E854”).

### Phenome-wide association analysis

To identify diagnoses associated with carrying the V122I variant, we performed a phenome-wide association analysis (PHEWAS) that included 427 ICD10 diagnosis codes in the unrelated Black UK Biobank subpopulation. After controlling for multiple comparisons, one diagnosis, polyneuropathy, was significantly associated with the V122I genotype (odds ratio [OR] = 6.4, 95% confidence interval [CI] = 2.6–15.6, *P* = 4.2 × 10^−5^) (Fig. 1; Supplementary Table 1). Significant associations between the V122I variant and polyneuropathy diagnosis were also seen in analyses of African Americans from the Penn Medicine Biobank (OR = 1.6, 95% CI = 1.2–2.4, *P* = 6.0 × 10^−3^) and the Million Veteran Program (OR = 1.5, 95% CI = 1.2–1.8, *P* = 1.8 × 10^−4^). In total, 2.1% (*n =* 5) of V122I carriers in the UK Biobank, 9.0% (*n =* 17) of V122I carriers in the Penn Medicine Biobank, and 4.8% (*n =* 111) of V122I carriers in the Million Veteran Program had a polyneuropathy diagnosis. The association of the V122I variant with polyneuropathy in the UK Biobank remained significant after adjustment for diabetes diagnosis (OR = 6.3, 95% CI = 2.6–15.4, *P* = 6.4 × 10^−5^).

**Fig. 1.**
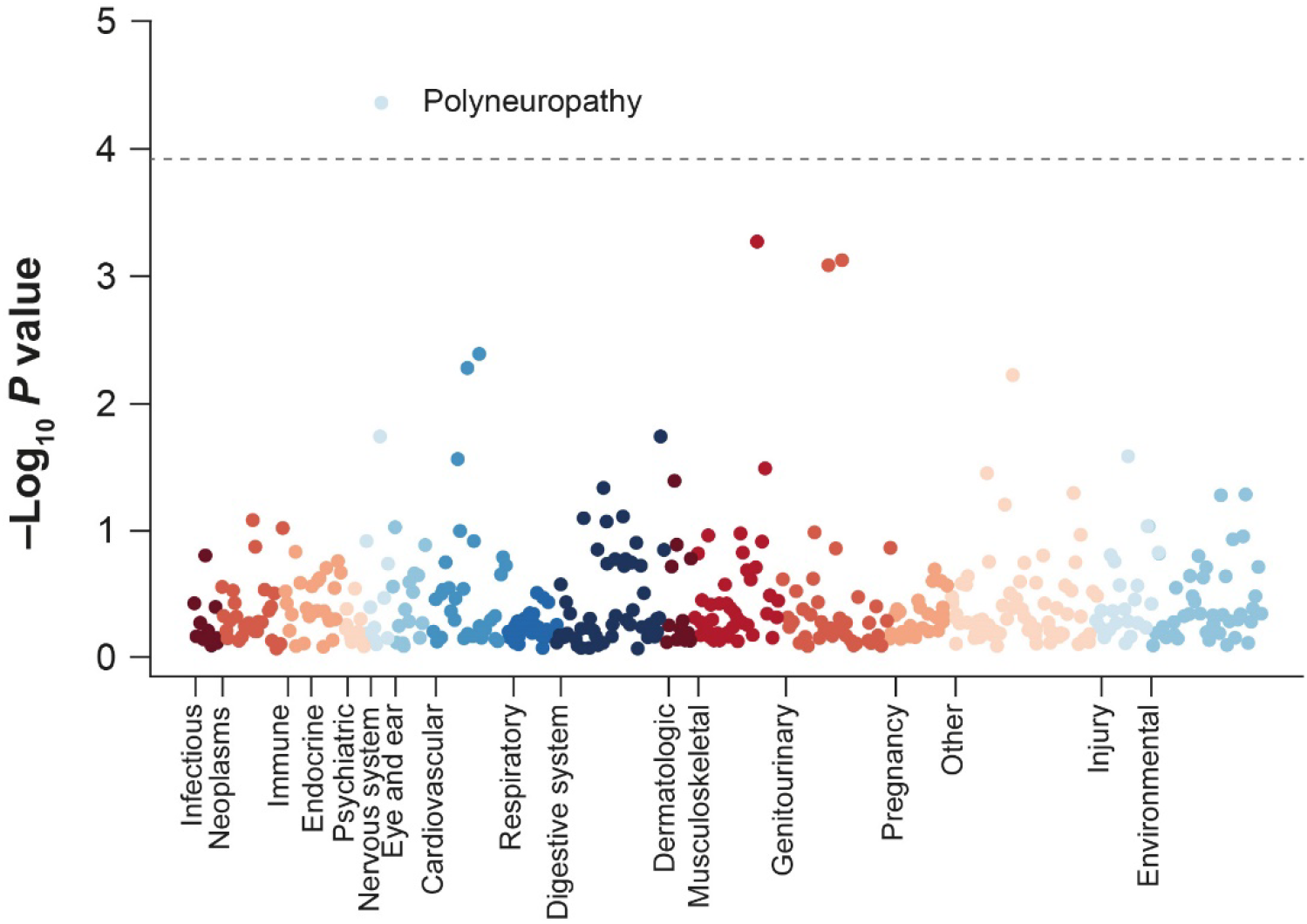
Phenome-wide association study of the V122I variant and 427 ICD10 diagnosis codes in the Black UK Biobank population. The dotted gray line indicates a multiple comparison-corrected significance cutoff of *P* < 1.2 × 10^−4^. *ICD10* International Statistical Classification of Diseases and Related Health Problems, 10th Revision, *V122I* valine-to-isoleucine substitution at position 122

### The V122I genotype and risk of common hATTR amyloidosis manifestations in the UK Biobank

In addition to unbiased PHEWAS analysis, we aimed to better understand the association between the V122I genotype and diagnosis of four common hATTR amyloidosis manifestations: polyneuropathy, CTS, cardiomyopathy, and HF, hereafter termed “common hATTR amyloidosis manifestations”. V122I carriers were significantly more likely to have at least one of these common diagnoses, with 11.1% of V122I carriers having at least one common hATTR amyloidosis manifestation compared with 4.9% of non-carriers (OR = 2.66, *P* = 1.5 × 10^− 6^). Prevalence of each of the four manifestations was higher in V122I carriers than non-carriers (Supplementary Fig. 3), and two of the three participants (66.7%) homozygous for V122I were diagnosed with at least one of the common hATTR amyloidosis manifestations. Significant associations between the V122I genotype and common hATTR amyloidosis manifestations were also observed in the Million Veteran Program (Supplementary Table 2).

We tested the time to first diagnosis of a common hATTR amyloidosis manifestation by V122I genotype using Cox proportional hazards regression in the UK Biobank, and found a significant association (hazard ratio [HR] = 2.77, 95% CI = 1.72–4.47, *P* = 2.62 × 10^−5^; Fig. 2). By age 65, the cumulative incidence of having at least one common hATTR amyloidosis manifestation among V122I carriers was 11.9% (95% CI = 3.1–19.8), which increased by age 75 to 37.4% (95% CI = 20.5–50.7) and was significantly higher than in non-carriers (13.8%, 95% CI = 11.6–16).

**Fig. 2.**
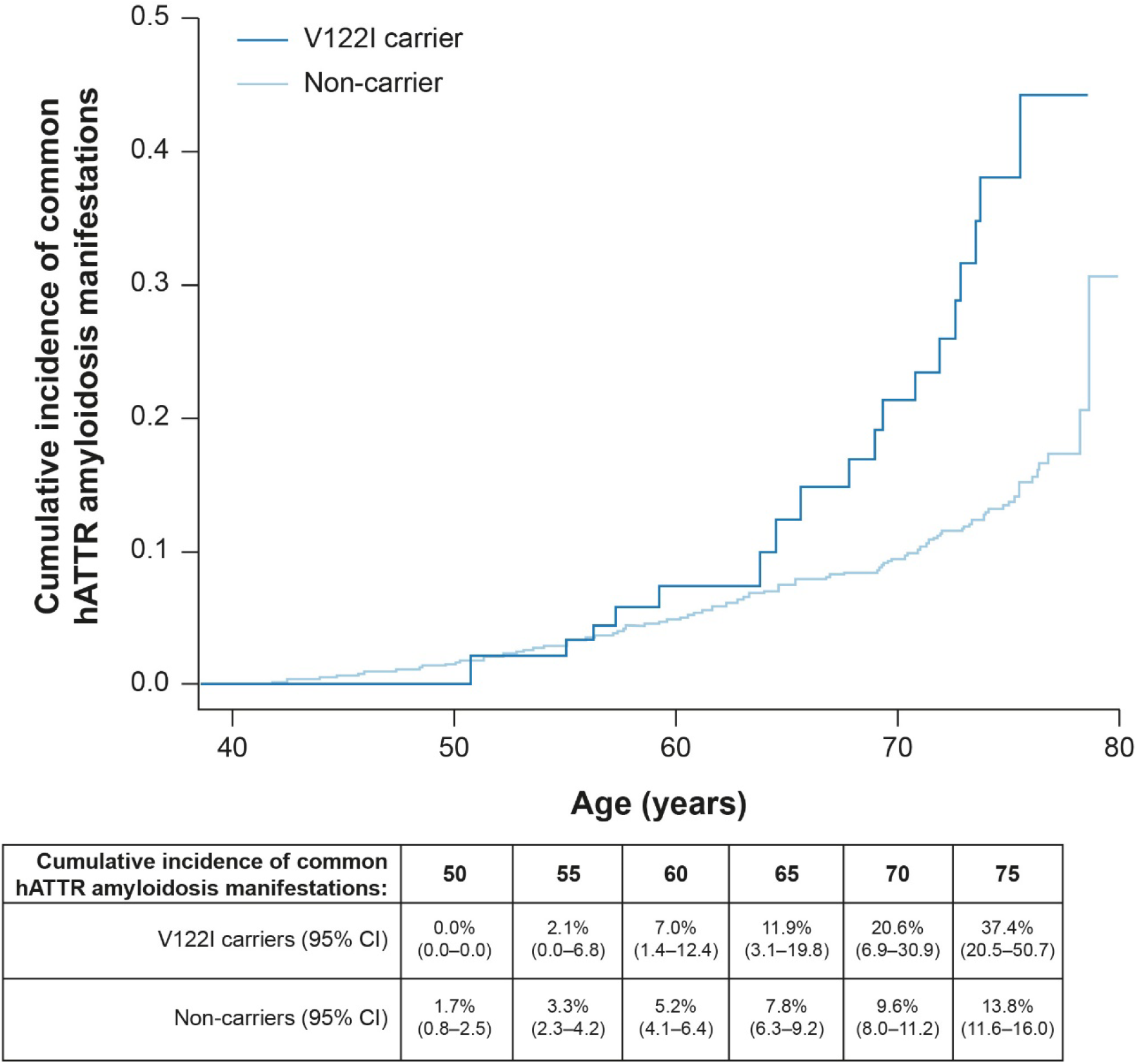
Cumulative incidence of common hATTR amyloidosis manifestations by V122I genotype in the UK Biobank (HR = 2.77, *P* = 2.62 × 10^−5^). *CI* confidence interval, *hATTR* hereditary transthyretin-mediated, *HR* hazard ratio, *V122I* valine-to-isoleucine substitution at position 122

The time to first common hATTR amyloidosis manifestation was assessed separately for each diagnosis (Fig. 3; Supplementary Table 3), finding that V122I carriers had a significantly higher cumulative incidence of polyneuropathy, CTS, and HF than non-carriers (polyneuropathy HR = 6.9, *P* = 9.5 × 10^−4^; CTS HR = 2.7, *P* = 0.01; cardiomyopathy HR = 3.2, *P* = 0.06; HF HR = 3.2, *P* = 2.0 × 10^−3^). By age 75, 7.9% of V122I carriers had a polyneuropathy diagnosis, 13.2% had a CTS diagnosis, 2.5% had a cardiomyopathy diagnosis, and 16.7% had an HF diagnosis. Cardiomyopathy and HF tended to be more prevalent in V122I carriers than non-carriers at older ages (Fig. 3c and d), whereas polyneuropathy was more prevalent at younger ages (Fig. 3a).

**Fig. 3.**
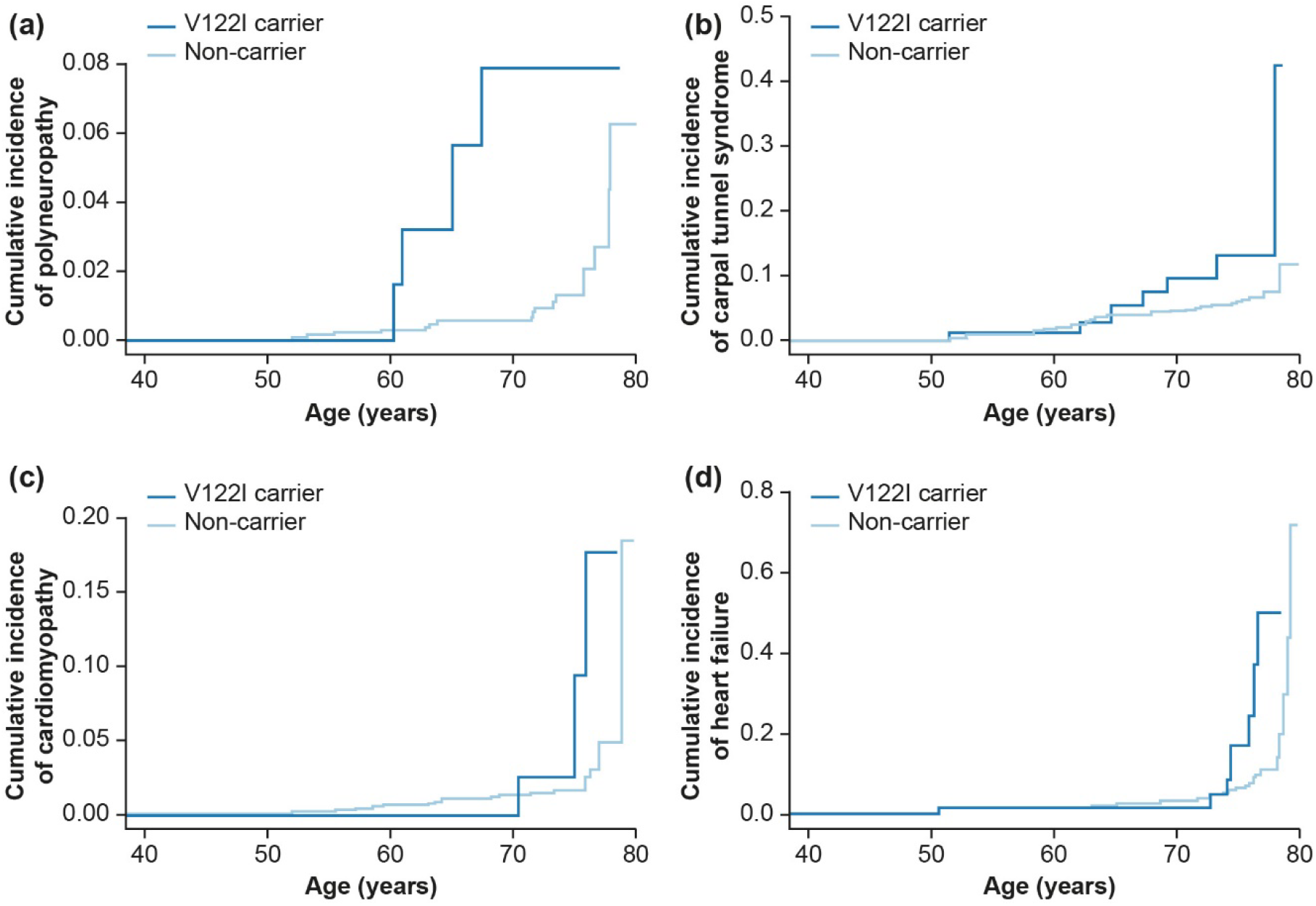
Cumulative incidence by V122I genotype of first diagnosis of common hereditary transthyretin-mediated amyloidosis manifestation: (**a**) polyneuropathy (G62), (**b**) carpal tunnel syndrome (G560), (**c**) cardiomyopathy (I42), and (**d**) heart failure (I50 or I098). *V122I* valine-to-isoleucine substitution at position 122

### Characteristics of V122I carriers with common hATTR amyloidosis manifestations in the UK Biobank

It remains unclear why some V122I carriers develop hATTR amyloidosis while others do not. To better understand the characteristics of V122I carriers with hATTR amyloidosis manifestations, we performed multiple logistic regression and demonstrated that V122I carriers with at least one common hATTR amyloidosis manifestation (*n =* 27) were more likely to be older and current/former cigarette smokers than those without common manifestations (*n =* 216). No sex difference was observed between those with and without any of the four common hATTR amyloidosis manifestations (*P* = 0.18) (Supplementary Table 4).

### Population attributable risk of common hATTR amyloidosis manifestations to the V122I variant

To assess the impact of the V122I variant on common hATTR amyloidosis manifestations in the UK Biobank Black population, we calculated the population attributable risk. Overall, 16.7% of the risk of a polyneuropathy diagnosis, 4.1% of the risk of a CTS diagnosis, 2.4% of the risk of a cardiomyopathy diagnosis, and 6.5% of the risk of an HF diagnosis were attributable to the V122I variant (Table 2).

**Table 2.** Attributable risk of common hATTR amyloidosis manifestations in the UK Biobank Black population (*n =* 6062) due to the V122I variant

| | Diagnoses in V122I carriers,<br>% ($n/N$) | Population attributable<br>risk to V122I variant, % |
| --- | --- | --- |
| Polyneuropathy | 20.0 (5/25) | 16.7 |
| CTS | 7.5 (13/174) | 4.1 |
| Cardiomyopathy | 7.5 (3/40) | 2.4 |
| HF | 10.2 (10/98) | 6.5 |
| At least one common hATTR amyloidosis manifestation | 8.7 (27/311) | 5.5 |
| *CTS* carpal tunnel syndrome, *hATTR* hereditary transthyretin-mediated, *HF* heart failure, *V122I* valine-to-isoleucine substitution at position 122 |  |  |

## Discussion

In this analysis of three large biobanks (UK Biobank, Penn Medicine Biobank, and the Million Veteran Program) including 2739 carriers, the V122I variant was significantly associated with a polyneuropathy diagnosis. This suggests that individuals with the V122I genotype, who were historically assumed to be predominantly at risk for cardiomyopathy, have a significantly increased risk of polyneuropathy. In the UK Biobank, by age 75, 7.9% of V122I carriers had a clinical diagnosis of polyneuropathy. Moreover, V122I carriers represent one in five of all polyneuropathy diagnoses in this subpopulation of the UK Biobank. The V122I genotype has been linked to polyneuropathy in case reports and smaller studies [22-24], as well as highlighted within a recent scientific statement from the American Heart Association [25]. However, to our knowledge, this is the first study to show a significant association between the V122I genotype and polyneuropathy. Additionally, an overall enrichment of common hATTR amyloidosis manifestations (polyneuropathy, CTS, cardiomyopathy, and HF) was found among V122I carriers. Despite this clear enrichment, only two out of 243 patients were diagnosed with amyloidosis (ICD10 “E85”), suggesting a considerable underdiagnosis of the disease that is consistent with previous literature [4, 5].

Notably, the two participants with confirmed amyloidosis were diagnosed in their 70s, with one dying from cardiac amyloid less than 2 years after diagnosis. This highlights the need for a better understanding of the early and multisystem manifestations of hATTR amyloidosis, particularly for V122I carriers who have higher mortality rates and worse prognosis than patients with other cardiac pathologies [9, 26-28].

To this end, the cumulative incidence of the four common hATTR amyloidosis manifestations was analyzed separately, finding that V122I carriers had a significantly higher cumulative incidence of polyneuropathy, CTS, and HF than non-carriers. Moreover, V122I carriers were more likely than non-carriers to have polyneuropathy at a younger age and cardiac manifestations at older ages. This finding implies that diagnosing polyneuropathy may be key to earlier disease identification.

In order to identify the characteristics of participants with common hATTR amyloidosis manifestations, differences between V122I carriers with and without any diagnoses of the common hATTR amyloidosis manifestations were analyzed. V122I carriers with these diagnoses were older and more likely to be current/former smokers. However, a difference between men and women in the prevalence of common hATTR amyloidosis manifestations was not found, despite previous studies identifying gender as a significant modifier of disease penetrance [29]. While the two amyloidosis diagnoses were both in males, diagnoses of common hATTR amyloidosis manifestations were present in equal proportions of males and females.

The results are particularly important given recent advances in therapies for hATTR amyloidosis. Two major strategies have been employed: 1) reduce serum TTR levels by inhibiting hepatic synthesis of TTR proteins through RNA interference therapeutics (patisiran) or antisense oligonucleotides (inotersen); and 2) prevent dissociation of the TTR tetramer through small molecule TTR stabilizers (tafamidis). Both approaches have yielded positive results in phase 3 studies of hATTR amyloidosis with polyneuropathy [30, 31], and tafamidis has also shown benefit in patients with ATTR amyloidosis with cardiomyopathy [32], with studies underway for patisiran [33] and inotersen [34]. For patients with hATTR amyloidosis to derive maximal benefit from the available therapies, early identification of patients remains essential.

Previous studies of V122I carriers have captured patients late in their disease course and/or looked at a limited set of predefined outcomes [35]. The main strength of this study is its size and prospective nature. Moreover, we identify treatable clinical manifestations associated with carrying a variant common in individuals of African ancestry, who are historically understudied in genetic research [36, 37]. However, this study must be interpreted in the context of potential limitations. Analyses in the UK Biobank were performed using inpatient hospital ICD10 diagnosis codes and do not capture outpatient or self-reported diagnoses. Therefore, it is possible our analyses missed additional hATTR amyloidosis manifestations present in participants but not captured in the hospital ICD10 diagnosis codes. Effect estimates of the association between the V122I variant and polyneuropathy were heterogeneous between the three biobanks (OR UK Biobank = 6.4; OR Penn Medicine Biobank = 1.6; OR Million Veteran Program = 1.5). This is likely due to differences in the collection methods of ICD diagnosis codes: in the UK Biobank, diagnoses were from inpatient hospital records, whereas in the Penn Medicine Biobank and the Million Veteran Program they included both inpatient and outpatient diagnoses. Indeed, the rate of polyneuropathy in non-V122I carriers is lower in the UK Biobank, suggesting that the higher OR in this population may be due to the method of ICD10 diagnosis collection.

Additional limitations include that UK Biobank participants are healthier on average than the general UK population [38]. Also, despite the large sample size, the number of V122I carriers with common hATTR amyloidosis manifestations in the UK Biobank is small (*n =* 27), limiting statistical power to detect significant associations. Furthermore, V122I carriers in the UK Biobank were on average 60 years of age at censoring (range: 42–79 years), which is younger than the age range during which V122I hATTR amyloidosis typically presents [35]. We hypothesize that this younger age may be a reason that the well-known cardiac manifestations of hATTR amyloidosis are not as enriched as expected based on previous reports. Lastly, while four common hATTR amyloidosis manifestations were found to be enriched in V122I carriers, not all diagnoses of cardiomyopathy, CTS, HF, and polyneuropathy can be attributed to hATTR amyloidosis, since these are commonly seen with other diseases as well as in an idiopathic manner.

In a large, unbiased study of one of the most common pathogenic *TTR* mutations, the V122I variant was significantly associated with polyneuropathy. This finding has important clinical implications, as physicians should have a high clinical suspicion for the multisystem manifestations of hATTR amyloidosis in V122I carriers, including both cardiomyopathy and polyneuropathy.

## Data Availability

UK Biobank data is available upon application to the UK Biobank.

## Supplementary Materials

### Materials and methods

#### Study populations

##### UK Biobank

The UK Biobank is a large, population-based, prospective cohort study which recruited 502,634 participants aged 40–69 years in England, Wales, and Scotland between 2006 and 2010 [1]. Participants in the UK Biobank lived within 25 miles of one of the 22 assessment centers and participated in a baseline assessment involving the collection of extensive data from questionnaires, health records, physical measurements, imaging, and biologic samples [1]. The UK Biobank recruited a population-based sample without selection on any disease outcome, but subjects were generally healthier than the overall National Health Service (NHS) population at baseline [2]. Data used in this analysis were accessed through UK Biobank application no. 26041 and are publicly available upon request to the UK Biobank. A total of 494,078 samples (including duplicates) were sent to Affymetrix for genotyping, with results from 488,377 included in the released data following quality control analysis [3].

All UK Biobank study participants underwent a baseline study assessment where covariates including age, sex, body mass index, self-reported diabetes diagnosis, self-reported hypertension diagnosis, and smoking status were collected. Additionally, participants consented to access to their NHS hospital records, which were used to obtain primary and secondary International Statistical Classification of Diseases and Related Health Problems, 10th Revision (ICD10) disease diagnosis codes. Diagnosis codes were collected prospectively from inpatient NHS visits during study follow-up and retrospectively from 1996 to baseline visit.

##### Penn Medicine Biobank

The Penn Medicine Biobank currently includes over 60,000 participants enrolled through the University of Pennsylvania Health System who have consented to allow the linkage of biospecimens to their longitudinal electronic health record (EHR). The current analysis utilizes data on 5737 individuals of genetically inferred African ancestry recruited between November 21, 2008 and January 4, 2017, that passed quality control as described in detail elsewhere [4].

##### Million Veteran Program

The Million Veteran Program is a large, multiethnic cohort within the U.S. Department of Veterans Affairs (VA). More than 825,000 veterans over age 18 years were recruited from 63 participating VA medical center facilities across the United States between 2011 and 2017 [5]. The Million Veteran Program biobank incorporates data from biospecimens, baseline and lifestyle surveys, and EHRs, including clinical laboratory measurements, diagnostic imaging reports, diagnosis and procedure codes, and vital status. Information from Medicare claims is not included in the EHR data. Million Veteran Program research protocols were approved by the VA Central Institutional Review Board. All participants provided informed consent and authorization for review of their medical records.

#### Phenome-wide association analysis and replication

Phenome-wide association analysis was performed in PLINK (v2.0) [6] using logistic regression with the “firth-fallback” option, which runs Firth regression [7] when logistic regression fails due to a rare outcome. Analyses were performed using the REVEAL/SciDB translational analytics platform from Paradigm4. The significant association between the valine-to-isoleucine substitution at position 122 (V122I) variant and polyneuropathy diagnosis was replicated in the Penn Medicine Biobank and the Million Veteran Program. In the Penn Medicine Biobank, polyneuropathy was defined as the assignment of the ICD9 diagnosis code 357 or ICD10 diagnosis codes of G62 or G63 on two or more separate dates. Association was assessed using logistic regression, controlling for age, sex, and the first five principal components. In the Million Veteran Program, polyneuropathy was defined as a diagnosis of “G62,” including child codes. Association was assessed using logistic regression, controlling for age, sex, and the first 10 principal components of genetic ancestry in 82,362 unrelated Million Veteran Program participants of African ancestry.

#### Assessment of common hATTR amyloidosis manifestations in V122I carriers

Time to first diagnosis of common hereditary transthyretin-mediated (hATTR) amyloidosis manifestations (polyneuropathy [“G62”], carpal tunnel syndrome [“G560”], cardiomyopathy [“I42”], or heart failure [“I50” or “I098”]) was tested in the UK Biobank unrelated Black population. For time to event analyses, prevalent diagnoses occurring before a patient’s date of enrollment were removed. Time on study was calculated as date of enrollment to date of first primary or secondary diagnosis of an hATTR amyloidosis manifestation (if participant had a diagnosis), date of lost to follow-up (if participant was lost to follow-up), or date of administrative censoring (March 31, 2017 for participants in England, February 29, 2016 for participants in Wales, or October 31, 2016 for participants in Scotland). Age at diagnosis was calculated from month and year of birth. Data availability dates were obtained from: http://biobank.ctsu.ox.ac.uk/crystal/exinfo.cgi?src=Data_providers_and_dates (accessed on April 1, 2019). Survival analyses were performed in R version 3.4.4 using the survival (v. 2.43) and survminer packages (v. 0.4.3). All analyses were controlled for known confounders including age, sex, smoking status, and genetic ancestry via 10 genetic principal components.

#### Population attributable risk to the V122I variant

The population attributable risk of common hATTR amyloidosis manifestations due to the V122I variant was calculated in the unrelated Black subpopulation of the UK Biobank (*n =* 6062). The following equation was used, where risk was estimated using the cumulative incidence of disease at age 75 from the Kaplan–Meier curve:

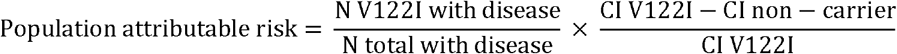

*CI* cumulative incidence

## Declarations

### Ethics approval

The UK Biobank study was approved by the National Health Service National Research Ethics Service (ref. 11/NW/0382) and all participants provided written informed consent to participate in the UK Biobank study. Information about ethics oversight in the UK Biobank can be found at https://www.ukbiobank.ac.uk/ethics/. This research has been conducted using the UK Biobank resource, application no. 26041. The Penn Medicine Biobank research was approved by the Institutional Review Boards of the University of Pennsylvania (Protocol nos.: 808346 [CGI], 813913 [PMBB], and 817977 [PMBB tissue]). The Million Veterans Program research was approved by the Institutional Review Boards of the Veterans Affairs (Protocol no. 19-06 – Genetics of Cardiometabolic Diseased in the VA Population II).

### Consent to participate

All necessary patient/participant consent has been obtained and the appropriate institutional forms have been archived.

### Consent for publication

Not applicable.

### Availability of data and material

UK Biobank data is available upon request to the UK Biobank. Data for the Penn Medicine Biobank and Million Veteran Program are available upon request.

### Code availability

Code written for this study is available upon reasonable request.

### Authors’ contributions

M.M.P., S.M.D., and C.T. had full access to the UK Biobank, Penn Medicine Biobank, and Million Veteran Program data analyzed in the study, respectively, and take responsibility for the integrity of the data and the accuracy of data analysis. K.F., A.K.V., G.H., P.N., and M.M.P. were involved in the concept and design. M.M.P., A.M.D., L.D.W., A.O.F-C., S.M.D., and C.T. were involved in the acquisition, analysis, or interpretation of data. All authors helped to interpret the data and critically revise the publication, are accountable for the accuracy and integrity of the publication, and provided final approval to submit for publication.

## Acknowledgment

This research has been conducted using the UK Biobank resource, application no. 26041. Funding was received from the U.S. Department of Veterans Affairs Office of Research and Development, Million Veteran Program (grant no. MVP003). This publication does not represent the views of the Department of Veterans Affairs or the U.S. Government. This research was also supported by additional Department of Veterans Affairs awards (no. I01-BX003362 to P.S.T. and K-M.C.) and used the resources and facilities at the Department of Veterans Affairs Informatics and Computing Infrastructure (no. VA HSR RES 13-457 to J.A.L.). S.M.D. is supported by the Veterans Administration (no. IK2-CX001780). Editorial assistance in the preparation of this manuscript was provided by Adelphi Communications Ltd, UK. The authors would like to thank Anastasia McManus, PharmD, RPh, of Alnylam Pharmaceuticals for her assistance in preparing this manuscript. The authors would also like to thank Paradigm4 for their assistance in conducting the phenome-wide association analysis.

---

**Supplementary Fig. 1.**
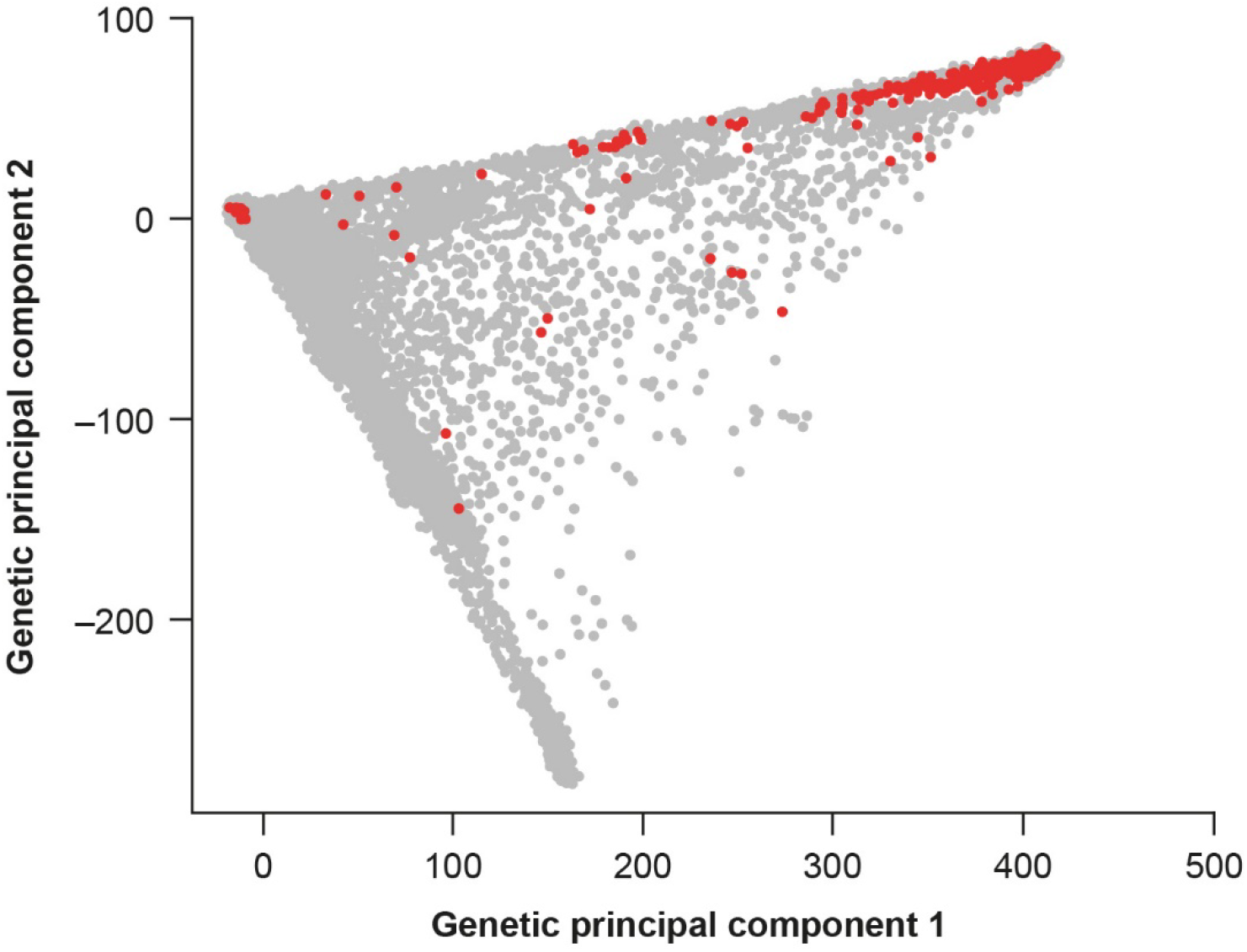
Genetic principal component 1 versus genetic principal component 2 with V122I carriers highlighted (red). The majority (80.6%) of V122I carriers are of African ancestry. The MAF of V122I in the four UK Biobank subpopulations was: 1) MAF Black subpopulation = 0.02; 2) MAF White subpopulation = 1.78 × 10^−5^; 3) MAF Southeast Asian subpopulation = 0.0; 4) MAF Chinese subpopulation = 0.0. *MAF* minor allele frequency, *V122I* valine-to-isoleucine substitution at position 122

**Supplementary Fig. 2.**
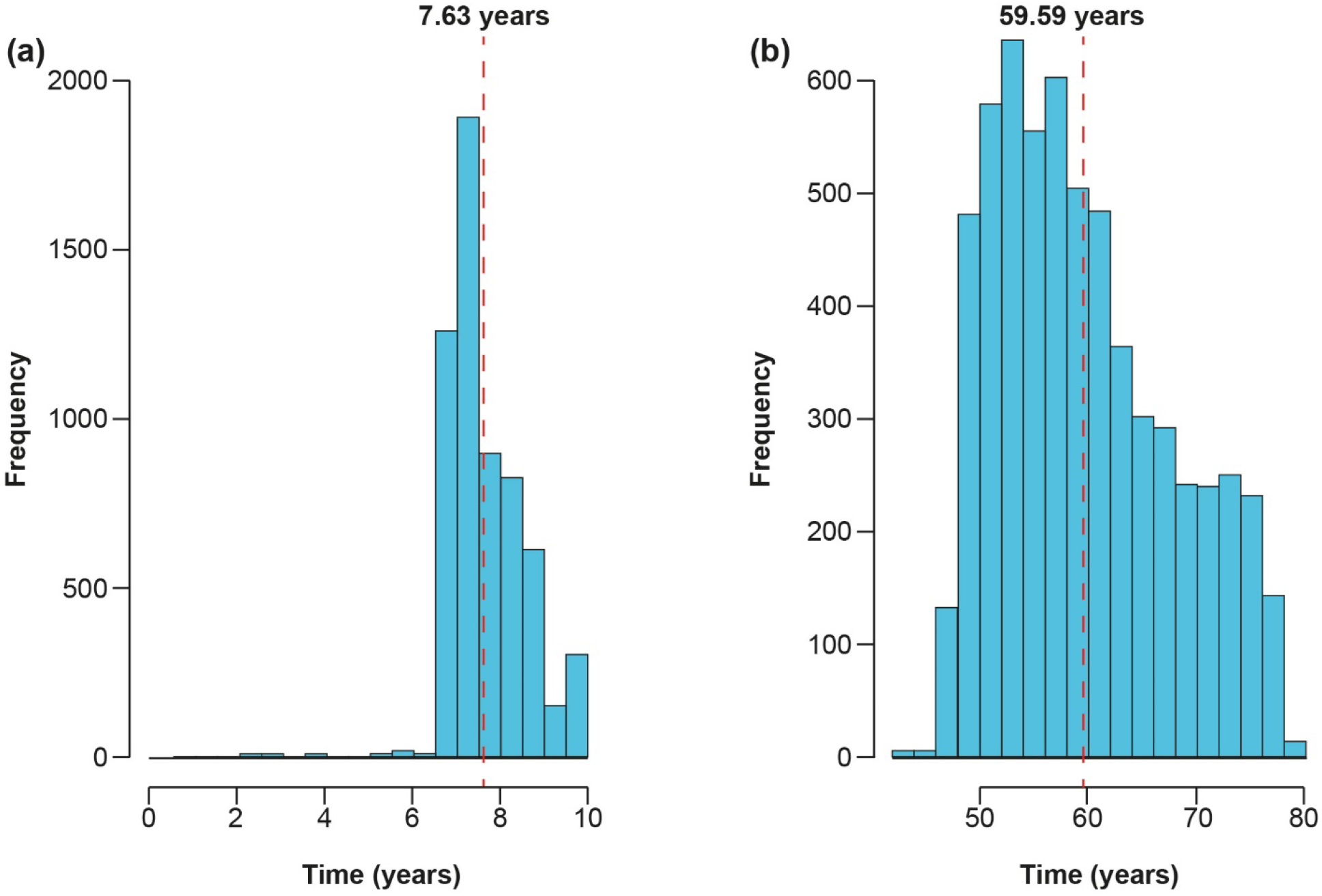
Histograms of (**a**) follow-up time in unrelated Black participants of the UK Biobank study (mean: 7.6 years, range: 0.22–10.0 years) and (**b**) age at last known UK Biobank observation (mean: 59.6 years, range: 42.2–79.0 years). Censoring occurred at date of death (if died), date of loss to follow-up (if lost to follow-up), or March 31, 2017 for participants in England, February 29, 2016 for participants in Wales, or October 31, 2016 for participants in Scotland. UK Biobank participants in this study were followed for a total of 46,260.8 years of person-time

**Supplementary Fig. 3.**
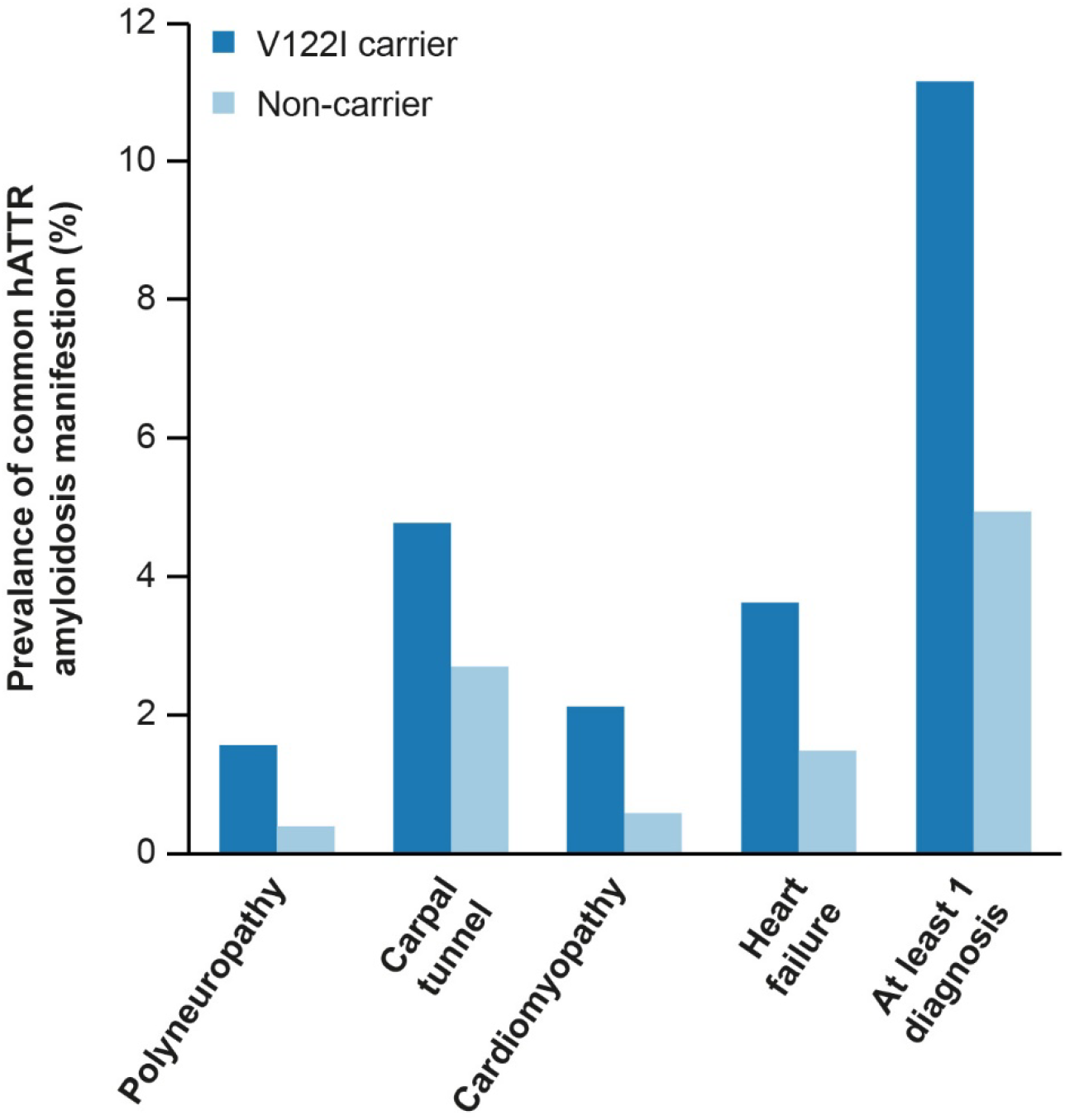
Prevalence of diagnoses frequently associated with hATTR amyloidosis in V122I carriers and non-carriers. *hATTR* hereditary transthyretin-mediated, *V122I* valine-to-isoleucine substitution at position 122

**Supplementary Table 1.** Top results from the phenome-wide association analysis of V122I variant and ICD10 diagnosis codes in the unrelated Black population of the UK Biobank

| ICD10 diagnosis code | Odds ratio | Standard error | *P* |
| --- | --- | --- | --- |
| Polyneuropathy (G62) | 6.40 | 0.45 | $4.24 \times 10^{-5}$ |
| Other disorders of synovium and tendon (M67) | 3.22 | 0.34 | $5.40 \times 10^{-4}$ |
| Unspecified lump in breast (N63) | 5.29 | 0.50 | $7.80 \times 10^{-4}$ |
| Epididymitis (N45) | 6.54 | 0.56 | $8.66 \times 10^{-4}$ |
| Heart failure (I50) | 2.50 | 0.32 | $4.35 \times 10^{-3}$ |
| Unspecified right bundle-branch block (I45) | 3.67 | 0.47 | $5.60 \times 10^{-3}$ |
| Retention of urine (R33) | 2.52 | 0.34 | $6.36 \times 10^{-3}$ |
| Mononeuropathies of upper limb (G56) | 1.94 | 0.29 | $1.98 \times 10^{-2}$ |
| Postprocedural disorders of digestive system (K91) | 4.20 | 0.62 | $2.03 \times 10^{-2}$ |
| Overexertion and strenuous or repetitive movements (X50) | 3.82 | 0.62 | $2.95 \times 10^{-2}$ |
| Non-rheumatic mitral valve insufficiency (I34) | 3.00 | 0.51 | $3.09 \times 10^{-2}$ |
| Other enthesopathies (M77) | 3.53 | 0.60 | $3.68 \times 10^{-2}$ |
| Ascites (R18) | 2.93 | 0.52 | $3.99 \times 10^{-2}$ |
| Other dermatitis (L30) | 3.33 | 0.60 | $4.63 \times 10^{-2}$ |
| Umbilical hernia (K42) | 2.04 | 0.37 | $5.17 \times 10^{-2}$ |

*ICD10* International Statistical Classification of Diseases and Related Health Problems, 10th Revision, *V122I* valine-to-isoleucine substitution at position 122

**Supplementary Table 2.** Association of the V122I variant with hATTR amyloidosis diagnosis and common hATTR amyloidosis manifestations in the Million Veteran Program cohort (*n =* 2306 V122I carriers and 80,076 non-carriers). Results are from a logistic regression controlling for age, sex, and genetic ancestry via the first 10 principal components

|  |  | Cases | Controls | Odds ratio | 95%<br>confidence<br>interval | *P* |
| --- | --- | --- | --- | --- | --- | --- |
| Amyloidosis | 869 | 81,513 | | 1.90 | 1.38–2.62 | $9.49 \times 10^{-5}$ |
| Cardiomyopathy | 3967 | 78,415 | | 1.34 | 1.12–1.61 | $1.42 \times 10^{-3}$ |
| Carpal tunnel syndrome | 4711 | 77,671 | | 1.69 | 1.45–1.97 | $1.95 \times 10^{-11}$ |
| Heart failure | 8717 | 73,665 | | 1.32 | 1.16–1.51 | $2.82 \times 10^{-5}$ |
| Polyneuropathy | 2745 | 79,637 | | 1.48 | 1.21–1.82 | $1.79 \times 10^{-4}$ |
| *hATTR* hereditary transthyretin-mediated, V122I valine-to-isoleucine substitution at position 122 |  |  |  |  |  |  |

**Supplementary Table 3.** Cumulative incidence of common hATTR amyloidosis manifestations by age. Estimates from Kaplan–Meier analysis testing time to first common hATTR amyloidosis manifestation by V122I genotype during UK Biobank study follow-up. Prevalent diagnoses occurring before a patient’s date of enrollment were removed. The association of V122I genotype with time to first common manifestation is tested using Cox proportional hazards regression controlling for age, sex, smoking, and genetic ancestry via 10 genetic principal components (polyneuropathy HR = 6.9, 95% CI = 2.2–21.7, *P* = 9.5 × 10^−4^; carpal tunnel syndrome HR = 2.7, 95% CI = 1.2–5.8, *P* = 0.01; cardiomyopathy HR = 3.2, 95% CI = 0.9–10.7, *P* = 0.06; heart failure HR = 3.2, 95% CI = 1.5–6.9, *P* = 0.002)

| Age | Cumulative incidence<br>V122I carriers (95% CI) | Cumulative incidence<br>non-carriers (95% CI) |
| --- | --- | --- |
| Polyneuropathy |  |  |
| 50 | 0.0% (0.0–0.0) | 0.0% (0.0–0.0) |
| 55 | 0.0% (0.0–0.0) | 0.2% (0.0–0.4) |
| 60 | 0.0% (0.0–0.0) | 0.3% (0.1–0.5) |
| 65 | 5.6% (0.0–11.8) | 0.6% (0.2–0.9) |
| 70 | 7.9% (0.0–15.2) | 0.6% (0.2–0.9) |
| 75 | 7.9% (0.0–15.2) | 1.3% (0.5–2.1) |
| Carpal tunnel syndrome |  |  |
| 50 | 0.0% (0.0–0.0) | 0.2% (0.0–0.3) |
| 55 | 1.2% (0.0–3.4) | 1.0% (0.6–1.4) |
| 60 | 1.2% (0.0–3.4) | 2.0% (1.4–2.6) |
| 65 | 5.4% (0.0–11.4) | 3.9% (3.0–4.9) |
| 70 | 9.9% (1.0–17.9) | 4.6% (3.5–5.7) |
| 75 | 13.2% (2.2–23.0) | 6.0% (4.6–7.5) |
| Cardiomyopathy |  |  |
| 50 | 0.0% (0.0–0.0) | 0.0% (0.0–0.0) |
| 55 | 0.0% (0.0–0.0) | 0.2% (0.0–0.4) |
| 60 | 0.0% (0.0–0.0) | 0.7% (0.3–1.0) |
| 65 | 0.0% (0.0–0.0) | 1.1% (0.6–1.7) |
| 70 | 0.0% (0.0–0.0) | 1.3% (0.7–2.0) |
| 75 | 2.5% (0.0–7.2) | 1.7% (0.9–2.4) |
| Heart failure |  |  |
| 50 | 0.0% (0.0–0.0) | 0.2% (0.0–0.3) |
| 55 | 1.1% (0.0–3.1) | 1.0% (0.6–1.4) |
| 60 | 1.1% (0.0–3.1) | 2.0% (1.4–2.6) |
| 65 | 1.1% (0.0–3.1) | 3.9% (3.0–4.9) |
| 70 | 1.1% (0.0–3.1) | 4.6% (3.5–5.7) |
| 75 | 16.7% (1.1–29.8) | 6.0% (4.6–7.5) |
| *CI* confidence interval, *hATTR* hereditary transthyretin-mediated, *HR* hazard ratio, *V122I* valine-to-isoleucine substitution at position 122 |  |  |

**Supplementary Table 4.** Characteristics of V122I carriers with ≥ 1 diagnosis of a common hATTR amyloidosis manifestation (polyneuropathy, carpal tunnel syndrome, cardiomyopathy, and heart failure)

| | No common manifestations ($n = 216$) | At least 1 common manifestation ($n = 27$) | *P* |
| --- | --- | --- | --- |
| Mean (SD) age, years | 51.70 (7.75) | 61.00 (7.80) | $2.81 \times 10^{-6}$ |
| Male, % | 47.70 | 37.00 | 0.18 |
| Mean (SD) BMI, kg/m<sup>2</sup> | 29.60 (5.03) | 30.90 (5.88) | 0.35 |
| Smoking, % | 25.50 | 40.70 | 0.014 |
| Genetic PC 1 | 384.00 (29.10) | 384.00 (33.10) | 0.89 |
| *BMI* body mass index, *hATTR* hereditary transthyretin-mediated, *PC* principal component, *SD* standard deviation, *V122I* valine-to-isoleucine substitution at position 122 |  |  |  |

